# Fallow time determination in dentistry using aerosol measurement

**DOI:** 10.1101/2021.01.26.21250482

**Authors:** Shakeel Shahdad, Annika Hindocha, Tulsi Patel, Neil Cagney, Jens-Dominik Mueller, Amine Kochid, Noha Seoudi, Claire Morgan, Padhraig Fleming, Ahmed Riaz Din

**Author notes:** **Role:** Contributed to conception, design, data acquisition and interpretation, drafted and critically revised the manuscript. **Role:** Design, data acquisition, drafted and critically revised the manuscript. **Role:** Contributed to conception, design, interpretation, drafted and critically revised the manuscript. **Role:** Contributed to conception, design, interpretation and critically revised the manuscript. **Role:** Contributed to conception and interpretation. **Role:** Contributed to conception and critically revised the manuscript. All authors gave their final approval and agree to be accountable for all aspects of the work.

## Abstract

**Aim:** To calculate fallow time (FT) required following dental aerosol generating procedures (AGPs) in both a dental hospital (mechanically ventilated) and primary care (non-mechanically ventilated). Secondary outcomes were to identify spread and persistence of aerosol in open clinics compared to closed surgeries (mechanically ventilated environment), and identify if extra-oral scavenging (EOS) reduces production of aerosol and FT.

**Methods:** In vitro simulation of fast handpiece (FHP) cavity preparations using a manikin was conducted in a mechanically and non-mechanically ventilated environment using Optical Particle Sizer™ and NanoScan™ at baseline, during the procedure and fallow period.

**Results:** AGPs carried out in the non-mechanically, non-ventilated environment failed to achieve baseline particle levels after one hour. In contrast, when windows were opened after AGP, there was an immediate reduction in all particle sizes.

In mechanically ventilated environments the baseline levels of particles were very low and particle count returned to baseline within 10 minutes following AGP. There was no detectable difference between particles in mechanically ventilated open bays and closed surgeries.

The effect of the EOS was greater in non-mechanically ventilated environment on reducing the particle count; additionally, it also reduced the spikes in particle counts in mechanically ventilated environments.

**Conclusion:** High-efficiency particulate air filtered mechanical ventilation along with mitigating factors (high-volume suction) resulted in reduction of FT (10 minutes). Non-ventilated rooms failed to reach baseline level even after one hour of FT. There was no difference in particle counts in open bay or closed surgeries in mechanically ventilated settings. The use of an EOS device can reduce the particulate spikes during procedures in both mechanical and non-mechanical environments.

This study confirms that AGPs are not recommended in dental surgeries where no ventilation is possible. No difference was demonstrated in FT required in open bays and closed surgeries in mechanically ventilated settings.

**Clinical significance:** AGPs should not be carried out in surgeries where ventilation is not possible. Mechanical ventilation for AGPs should be gold standard; where not available or practical then the use of natural ventilation with EOS helps reduce FT. AGPs can be carried out in open bay environment with a minimum of 6 air changes per hour of mechanical ventilation. Four-handed dentistry with high-volume suction and saliva ejector are essential mitigating factors during AGPs.

## Introduction

COVID-19 has the potential to spread during dental procedures through a number of routes. Attention has focused on the spread via droplets or ‘splatter’ that can either impact directly on the face of a susceptible person or, be deposited on a surface (Shahdad et al. 2020; Allison et al. 2021). However, there is increasing evidence that aerosols, particularly when highly concentrated in enclosed environments, may play an important role in disease transmission (WHO 2020; Li et al. 2020; Miller et al. 2020). Aerosols in dental procedures are typically defined as particles smaller than 5μm (WHO 2020) that can remain suspended in air for hours.

There have been extensive investigations attempting to characterise the potential for infection from aerosol to occur in dentistry (Leggat and Kedjarune 2001; Harrel and Molinari 2004; Zemouri et al. 2017). Many researchers have focussed on measuring bioaerosol using cultures to quantify the amounts of bacteria or fungi deposited on surfaces. However, this approach relies on bioaerosols settling onto the surface and cannot account for the particles that remain suspended in the air or those removed through ventilation (Bentley et al. 1994; Teanpaisan et al. 2001; Chuang et al. 2014; Holloman et al. 2015; Al-Amad et al. 2017; Zemouri et al. 2020; Mirhoseini et al. 2021).

Other researchers have added dye or fluorescent marker to the water lines to examine the distribution of splatter and detect deposits as small as 1000 μm^2^ in area, although this dimension exceeds what is typically classed as an aerosol (Chiramana et al. 2013; Veena et al. 2015; Shahdad et al. 2020; Allison et al. 2021). Small particles (≤16 – 27μm) deposited on microscope slides have also been studied, however, this did not account for aerosols that did not settle during the experiment (Junevicius et al. 2005).

In the past decade, there have been a small number of studies using particle counters to directly sample the concentration of aerosols suspended in the air (Polednik 2014); although the focus has often been on the nanoparticles released from dental composite materials rather than on the potential spread of infection (Bradna et al. 2017).

As a result of risks associated with COVID-19, routine use of fallow time (FT) to allow for settling of suspended aerosol has been recommended following aerosol-generating procedures (AGP). Routine adoption of FTs may limit the capacity for the provision of dental care. However, there has been little consistency in the definition of an AGP or indeed on, the necessity for and duration of FT following AGPs (Din et al. 2020; National Services Scotland Short Life Working Group 2020). Due to the lack of experimental data on aerosols produced during dental procedures, there is no consensus on the FT required after AGPs. Previous guidance from Public Health England (PHE Oct 2020) based on the New and Emerging Respiratory Virus Threats Advisory Group (NERVTAG), recommended a FT of 60 minutes in a single room with 6 air changes per hour (ACH) following AGPs (FGDP 2020; SDCEP 2020). A recent rapid review of international dental guidance documents found that most guidance’s did not refer to a FT. When FT was recommended this varied from between 2 and 180 minutes (Clarkson et al. 2020). The median FT was 15 minutes for “non-COVID-19” patients and 20 minutes for confirmed or suspected COVID-19 patients (Clarkson et al. 2020).

There has been an increase in availability of extraoral scavenger (suction) devices (EOS) on the market since the beginning of the COVID-19 pandemic. In a recently published study, EOS reduced the mean intensity of contamination and frequency of splatter during most of the simulated procedures in both open clinic and closed surgery in a dental hospital with mechanical ventilation at six air changes per hour (Din et al. 2020; Shahdad et al. 2020). However, the effect of EOS on aerosol after AGPs is yet to be proven. Therefore, the primary aim of this study was to calculate the FT required for aerosols produced during various simulated AGPs to return to baseline levels in a dental hospital (mechanical ventilation) and a primary care setting (non-mechanical ventilation).

Secondary aims were to i) to identify if an EOS reduces the production of aerosol and FT required following AGPs in dentistry and, ii) identify if spread and persistence of aerosol generated in an open clinic was worse than a closed surgery in a mechanically ventilated environment.

## Methodology

AGPs were simulated on a dental manikin in a multi-chair open clinic and closed surgery in a dental teaching hospital (The Royal London Dental Hospital, London UK). During all procedures, mechanical ventilation with six air changes per hour involving a centralised air exchange system remained operational. The procedures were repeated in a private dental clinic (Specialist Dental Services, 94 Harley Street, London, UK) with and without natural (non-mechanical) ventilation.

AGPs were carried out using a protocol using an air turbine (W&H Synea™ Turbine TA-98LED, Bürmoos, Austria) for 20 minutes while simulating cavity preparation of LL6, crown preparation of LL1 and UL1 on thermoplastic teeth. Handpieces were operated at approximately 360,000 RPM with air and water coolant at maximum flow.

All procedures were carried out using a four-hand dentistry technique which included the assistant operating high volume suction (HVS) and a saliva ejector (SE). The procedures were repeated three times by the same operators to reduce performance bias.

The procedure was repeated with EOS which was placed at the 5 o’clock position, 15cm from the mouth of the manikin and operated at maximum flow rate (TM10, TopMed Dental Lighting Co. Ltd, Foshan China; specified flow rate 310m3/h).

Aerosol measurement with particle assessment was undertaken using both an Optical Particle Scanner (Optical Particle Sizer 3330, TSI Inc Minnesota, USA), which measured particles in the range 0.3 – 10 μm, and a Spectrometer Particle Scanner (NanoScan SMPS Nanoparticle Sizer 3910, TSI Inc Minnesota, USA) which spanned the range 10nm –0.365μm, and operated as described previously (Din et al 2020). Briefly, the sampling inlets were placed adjacent to the manikin’s mouth in the 7 o’clock position (8cm from the UL1) for all procedures (Figure 1). Using both counters it was possible to measure particles in 26 bins, ranging from 10nm to 10μm in diameter. As a single SAR-CoV-2 virion is approximately 80 to 100 nm in diameter, formation of particles smaller than 80nm was deemed to be irrelevant to virus transmission and was discarded. In order to reduce the remaining dataset, the particle counts were combined into four categories: ‘very small’ (0.08 - 0.26 μm), ‘small’ (0.27 - 0.90 μm), ‘medium’ (0.91 - 2.70 μm) and ‘large’ (2.71 - 10 μm).

**Figure 1.**
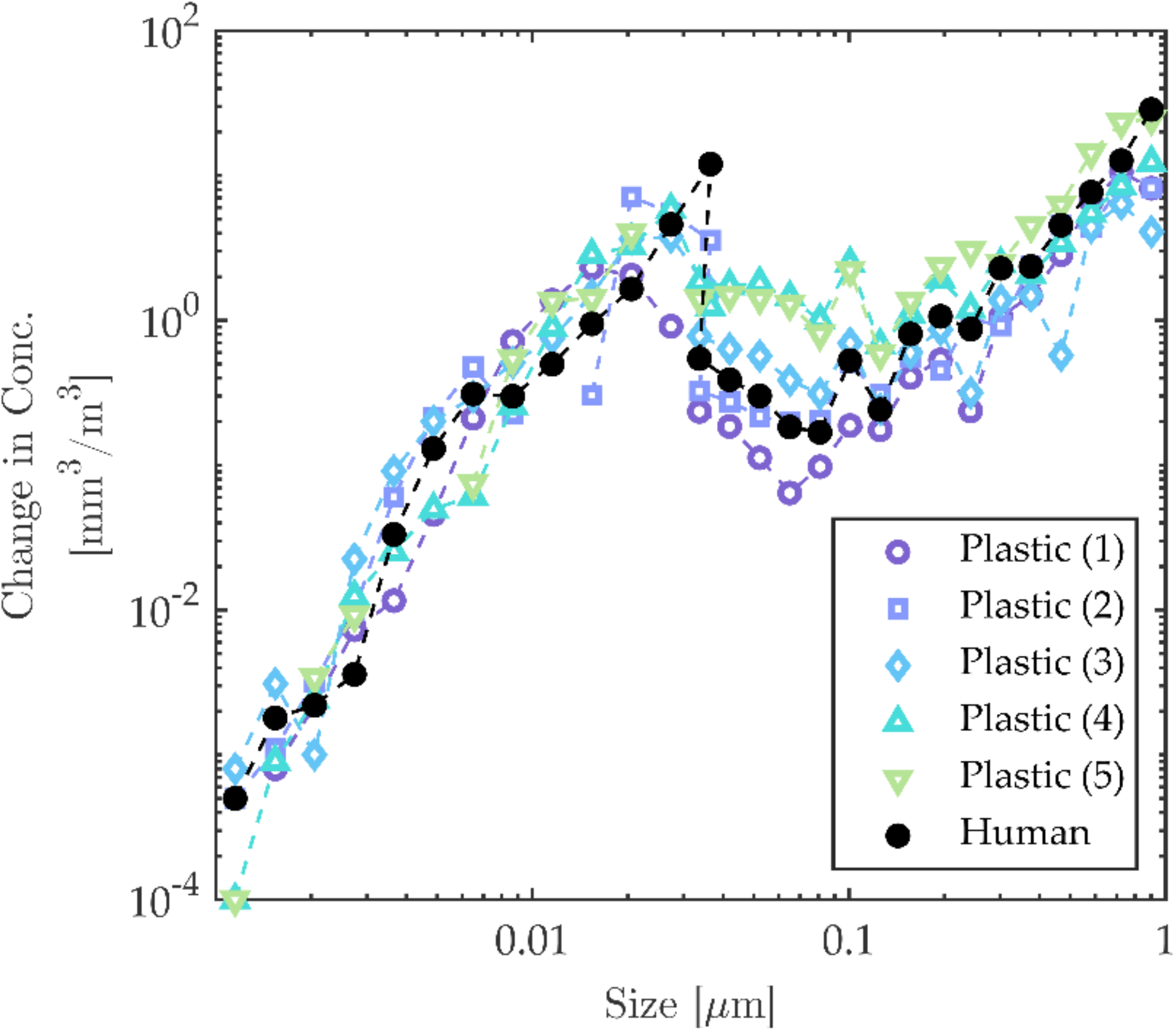
Optical Particle Sizer 3330 (TSI Inc Minnesota, USA) (on dental chair) and NanoScan SMPS Nanoparticle Sizer 3910 (TSI Inc Minnesota, USA) (on floor). The sampling inlets placed adjacent to the manikin’s mouth in the 7 o’clock position (8cm from the UL1)

Measurements were recorded continuously and each sample cycle took 1 minute to complete (Aerosol Instrument Manager 10.3.1.0 and NanoScan Manager 1.0.0.19, TSI Incorporated, Minnesota, USA). All experiments included a pre-operative measurement of ten minutes to allow for a baseline atmosphere or characteristics recording, followed by the procedure and finally by a post-procedure FT. The post-procedure FT was initially chosen as 60 minutes. However, pilot results indicated that this could be reduced to 30 minutes for procedures carried out in the mechanically ventilated setting.

For procedures involving the EOS, the pre-procedure particle measurement was recorded with 10 minutes of no activity followed by 10 minutes with the EOS functioning alone to evaluate the effect of EOS on baseline measurements. This was then followed by the AGP and the post-procedure FT as above.

Only the operator and assistant were present in the room prior to the procedure and they left immediately after completion of the procedure. The door was kept shut in the closed surgery during all times. In EOS procedures, the device was left functioning during the post-procedure FT. The operator and assistant wore fluid resistant surgical masks (FRSM) during the procedures and talking was allowed pre-procedure to simulate a typical dental appointment.

In order to study the spread of aerosol, open clinic experiments were repeated with the particle-measuring equipment positioned at varying distances from the manikin including an adjacent bay (at a distance of 1.7m and over a partition wall measuring 1.2m tall) and opposite bays (at a distance of 1.7m with no intersecting partition).

In order to identify the aerosols actually produced by a fast handpiece (FHP), the minimum concentration of aerosols found for each size in the pre-procedural 10-minute period was subtracted from the corresponding median value found during the procedure.

For purpose of external validity, the experiment was repeated to measure size distribution of aerosol particles using FHP for 7 minutes on human extracted teeth and compared with plastic teeth under identical conditions in the non-mechanically ventilated environment with closed windows.

### Statistics

Descriptive analysis was used to identify the characteristic in aerosol change in the various particle size groups; these were then represented visually in frequency graphs showing concentration (mm^3^/m^3^) over time (minutes) with the procedure period highlighted in yellow.

Further statistical analysis included the calculation of the post-procedure FT and this was represented by the time set at when the particle concentration (in each particle size group) reaching a threshold within a 5% of the mean of the pre-procedural particle concentration.

A Wilcoxon rank sum test was used to assess whether the EOS resulted in a reduction in the median overall FT for the closed and open bay cases in the hospital setting.

## Results

### Aerosol Generation & Effect of Extra-Oral Suction

Figure 2 summarises the aerosols measured throughout each of the experiments. These involved the use of FHP for 20 minutes simulating the same procedure but under a variety of clinical conditions (yellow shaded region), both without (blue) and with (red) use of the EOS device.

**Figure 2.**
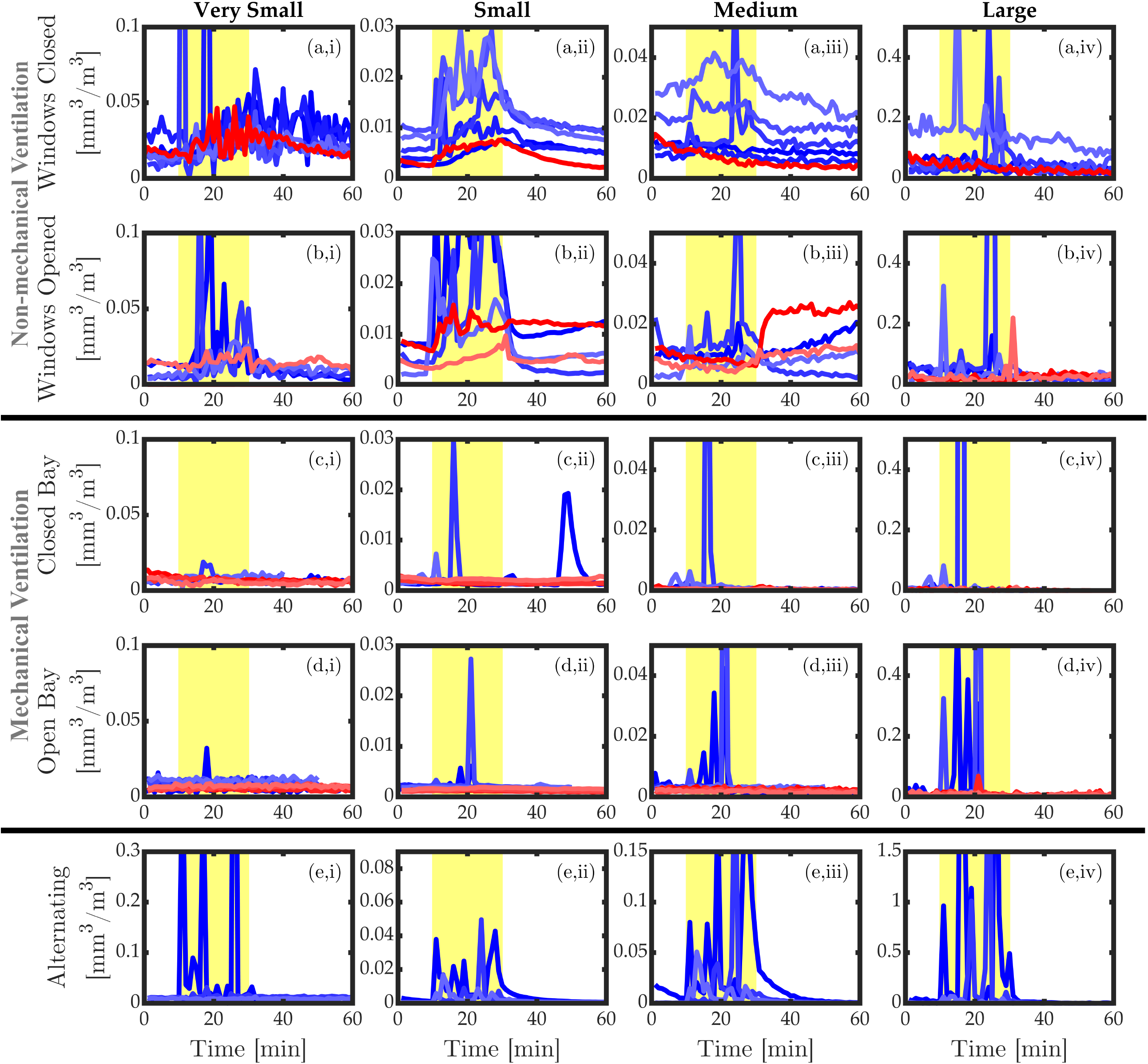
Variation in measured aerosol levels for the same procedure (20 minutes drilling using FHP) repeated under different conditions; The blue lines represent recordings without EOS and red lines denote those including EOS. The columns represent the different size ranges of particles; ‘very small’ (i, 0.08 - 0.26 μm), ‘small’ (ii, 0.27 - 0.90 μm), ‘medium’ (iii, 0.91 - 2.70 μm) and ‘large’ (iv, 2.71 -10 μm). The rows correspond to different surgery set-up; non-mechanically ventilated environment with closed windows (a), non-mechanically ventilated environment with windows opened at end of the procedure (b), mechanically ventilated environment in a hospital closed surgery (c), open hospital clinic with mechanically ventilated environment (d), in which the tooth being drilled was alternated every 5 minutes (e). Yellow shaded regions indicate the duration of the procedure; the time preceding represents the initial pre-treatment period and the time after represents the post-treatment FT. (N.B. the limits of the y-axes in (e) are three times higher than those of other rows).

Aerosol levels were highest and most sustained in non-mechanically ventilated environment with the windows closed throughout (Figure 2a). There was a distinct increase in the concentration of aerosols across the ‘very small’, ‘small’ and ‘medium’ ranges during the procedure. In some cases, this increase occurred in the form of large, isolated spikes that arose apparently randomly (Figure 2a,i) at 10 minutes and Figure 2(a,iii) at 32 minutes. Apart from these spikes, there was a clear tendency for the concentration to gradually increase throughout the procedure, and then slowly decline afterwards. This was most apparent in the ‘small’ size range (Figure 2(a,ii)), where in some cases the concentration did not return to baseline levels up to 90 minutes after the procedure finished (Table 1).

**Table 1.** Estimated FT (in minutes) required for aerosol levels in each particle size range to return to within 5% of their initial concentration, for each experiment. The right most column shows the largest FT identified in each experiment. In some cases, the aerosol levels never returned to within 5% of their original level; these cases are denoted as ‘>X’, where X is the time taken until the end of the measurement.

| Procedure | Number | Very small | Small | Medium | Large | Overall $\Sigma$(min) |
| --- | --- | --- | --- | --- | --- | --- |
|  |  | (0.08–0.26 µm) | (0.27–0.90 µm) | (0.91–2.60 µm) | (2.61–10 µm) |  |
| Windows Closed | 1 | 17 | >30 | 1 | 1 | >30 |
|  | 2 | 1 | >41 | 1 | 2 | >41 |
|  | 3 | 8 | >60 | 8 | 16 | >60 |
|  | 4 | 13 | 27 | 10 | 2 | 27 |
|  | 5 | 36 | 31 | 12 | 1 | 36 |
|  | Median |  |  |  |  | >30 |
|  | Max |  |  |  |  | >60 |
| Windows Closed (with | 1 | 2 | 24 | 1 | 1 | 24 |
| EOS) | **Median** |  |  |  |  | **24** |
|  | **Max** |  |  |  |  | **24** |
| Closed Bay | 1 | 1 | 1 | 8 | 3 | 8 |
|  | 2 | 2 | 1 | 6 | 1 | 6 |
|  | 3 | 2 | 7 | 4 | 1 | 7 |
|  | **Median** |  |  |  |  | **7** |
|  | **Max** |  |  |  |  | **8** |
| Closed Bay (with EOS) | 1 | 1 | 1 | 3 | 1 | 3 |
|  | 2 | 1 | 1 | 4 | 2 | 4 |
|  | 3 | 1 | 1 | 5 | 5 | 5 |
|  | **Median** |  |  |  |  | **4** |
|  | **Max** |  |  |  |  | **5** |
| Open Bay | 1 | 1 | 1 | 2 | 7 | 7 |
|  | 2 | 17 | 3 | 2 | 2 | 17 |
|  | 3 | 1 | 1 | 1 | 1 | 1 |
|  | 4 | 5 | 1 | 1 | 1 | 5 |
|  | 5 | 2 | 1 | 2 | 1 | 2 |
|  | **Median** |  |  |  |  | **5** |
|  | **Max** |  |  |  |  | **17** |
| Open Bay (with EOS) | 1 | 2 | 1 | 1 | 1 | 2 |
|  | 2 | 1 | 1 | 1 | 2 | 2 |
|  | 3 | 24 | 1 | 1 | 1 | 24 |
|  | 4 | 1 | 2 | 1 | 1 | 2 |
|  | **Median** |  |  |  |  | **2** |
|  | **Max** |  |  |  |  | **24** |
| Alternating handpiece direction | 1 | 2 | 12 | 8 | 3 | 12 |
|  | 2 | 6 | >30 | 26 | 6 | >30 |
|  | 3 | 1 | 26 | 18 | 1 | 26 |
|  | **Median** |  |  |  |  | **26** |
|  | **Max** |  |  |  |  | **>30** |

When the EOS was used (red), the magnitude of the increase in aerosol particles during the procedure was reduced. The signals contain fewer large spikes and appeared to take less time to revert to baseline levels. In some cases (the ‘medium’ and ‘large’ size ranges), the use of EOS resulted in a continuous reduction in aerosol levels throughout the entire experiment, indicating that the EOS was filtering out other background aerosols, as well as those produced by the operative procedure.

The equivalent data with the windows opened immediately following the end of the procedure (at 30 minutes) and left open during the whole post-procedure FT in the non-mechanically ventilated environment is illustrated in Figure 2(b). During the procedure, the use of the EOS device led to lower aerosol levels. In the case of the ‘very small’ and ‘small’ particles, the opening of the window coincided with a sudden reduction in aerosol levels, while in one case there was a sharp increase (Figure 2b,iii).

When the procedures were repeated in the mechanically ventilated environment of the dental hospital, there was a distinctly different pattern. The closed bay of (approximately 6 ACH) exhibited lower pre-procedural baseline particle levels, with notably fewer spikes and no appreciable increase in concentration over the course of the procedure (Figure 2(c)) compared with non-mechanically environment in the practice setting. No clear differences were observed for the test performed in the open bay in the mechanically ventilated environment of the hospital (Figure 2(d)).

The additive effect of EOS was less noticeable in the hospital within a HEPA filtered mechanical ventilation environment during the open and closed bay experiments, implying that the ventilation system was sufficiently effective. With the exception of a minor small spike at 20 minutes in one instance (Figure 2d,iv), no evident large particle spikes were recorded during the AGP when the EOS was used, suggesting that EOS could effectively prevent or reduce the frequency of high levels of particles generated by aerosol.

We hypothesised that these particle spikes occurred due to the relative proximity of the FHP to the intra-oral suction device inlets (HVS & SE) and the water spray was not being effectively removed when moved or repositioned in the mouth. In order to verify this effect, we repeated the FHP procedure without EOS, but changed the tooth that was being operated on every 5 minutes; alternating between the upper and lower anterior teeth Figure 2(e). A very large increase in aerosol levels at 5-minute intervals was observed corresponding to the change in FHP position. It should be noted that the limits of the y axes in Figure 2(e) are three times greater than the corresponding graphs discussed previously. This confirmed that brief changes in the position of the FHP, HVS or SE can lead to the release of large amount of aerosol into the near vicinity.

### Open Bay Procedures

In order to further investigate the behaviour of aerosols released during AGPs in open bay clinics, the procedure was repeated a number of times with the particle counters placed at adjacent and opposite bays. This effectively measured the potential for aerosol generated at one bay to lead to transmission to a patient or practitioner in a nearby bay. The time-series for the various particle size ranges are shown in Figure 2 for measurements performed on the operative chair, at an adjacent and opposite bay.

A series of spikes in aerosol concentration were observed during the procedure near the patient’s mouth. A minimal increase in medium particle size was observed in the adjacent bay. It is important to note the concentration is extremely low (y-axis) compared to the spike concentration on the patient.

### Tests with Human Teeth

The size distribution of aerosol particles measured over 7 minutes of operating on both human and plastic teeth under identical conditions (in a private practice with closed windows) was also assessed (Figure 3). In order to identify the aerosols actually produced by the FHP, the minimum concentration of aerosols found for each size in the pre-procedural 10-minute fallow period was subtracted from the corresponding median value found during the procedure. Little difference in the concentration with plastic and human extracted tooth tissue was observed (Figure 3).

**Figure 3.**
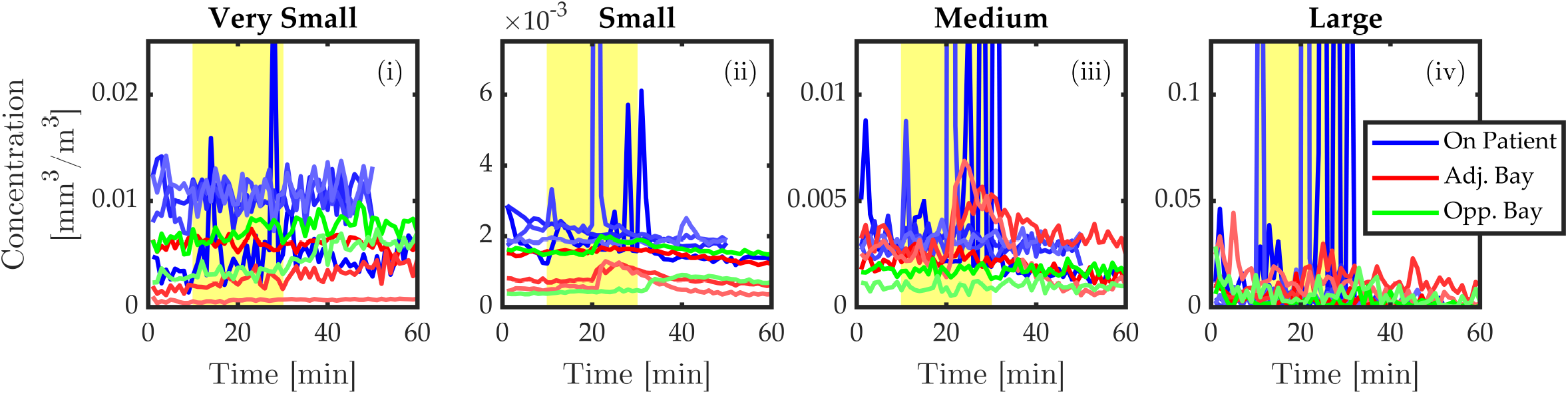
Variation in measured aerosol levels for the same procedure (20 minutes FHP) drilling using FHP) performed in an open bay with additional measurements taken in the opposite and adjacent bays., measured at various positions (with EOS). The columns represent the different size ranges of particles; ‘very small’ (i, 0.08 - 0.26 μm), ‘small’ (ii, 0.27 - 0.90 μm), ‘medium’ (iii, 0.91 - 2.70 μm) and ‘large’ (iv, 2.71 - 10 μm).

**Figure 4.**
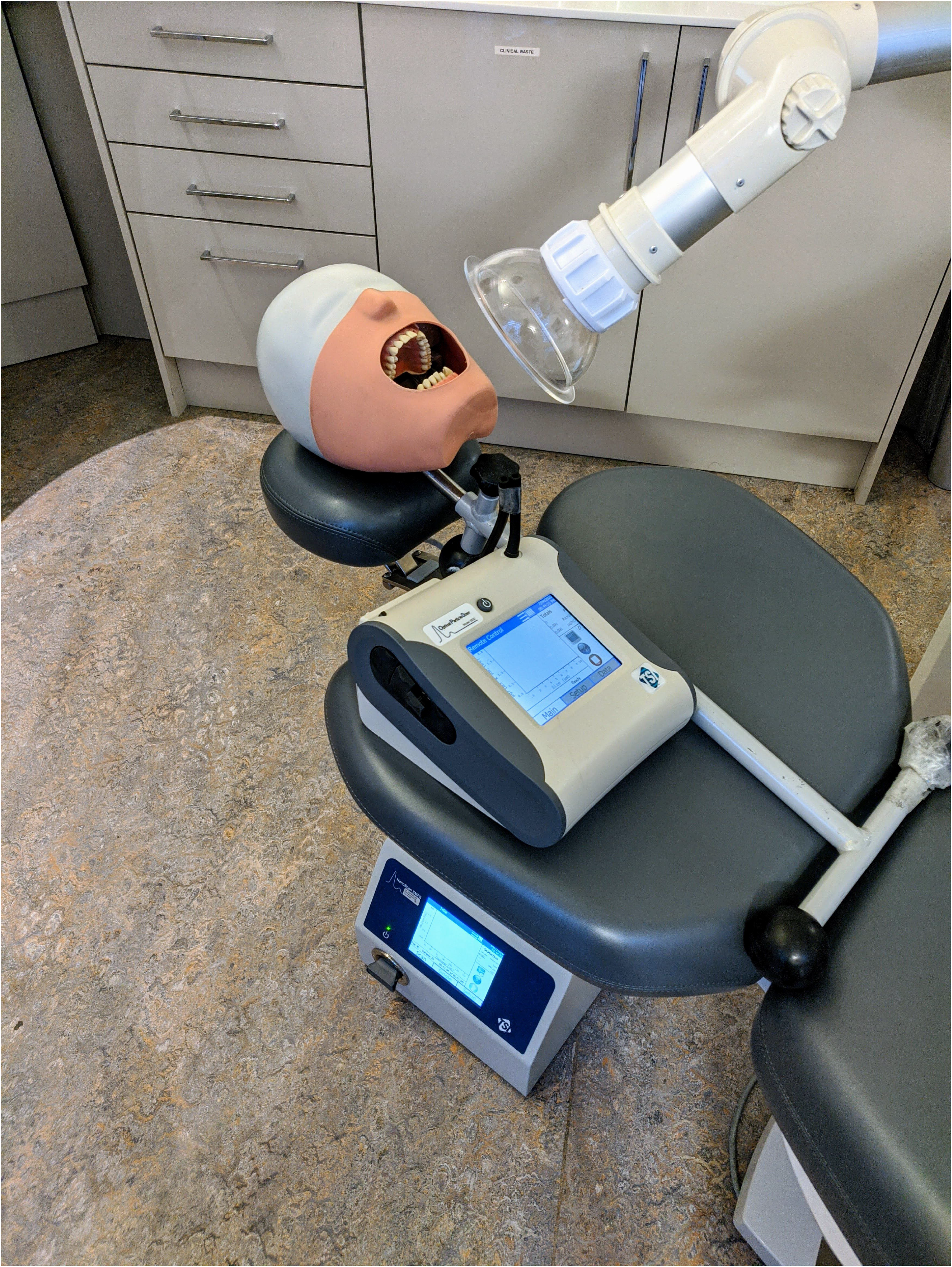
Change in concentration of aerosol levels, recorded over 7 minutes of drilling using FHP, relative to the minimum found in the 10 minutes pre-procedural time. The black symbols denote tests done using human tooth, while the other symbols correspond to repeated test performed using plastic teeth (the results of these tests are also shown in Figure 2(a)).

### Fallow Time Calculation

A key question when considering how dentistry can safely be resumed during the pandemic is how much FT is required at the end of a given procedure. This was estimated from the aerosol measurements (Figure 2) by calculating how long it took from the end of the procedure for the aerosol concentration in each size range to revert to within a threshold of 5% of the mean value before the procedure. A conservative approach was adopted with the overall FT taken as the longest identified for each particle size range (Table 1). With the exception of two cases, the FT estimates differed by less than 3 minutes when threshold values of 0.05% and 7.5% were used (with 15 out of 23 cases showing no change), indicating that these estimates were not sensitive to the threshold value chosen.

This method was not applied to the experiments in which the windows were opened at the end of the procedure because in these cases the change in the aerosol levels measured were reflective of those outside the window, and in some cases the air exchange led to a significant increase in the concentration (e.g. Figure 2b,iii). Therefore, increases in aerosol concentration post-procedure in these experiments would not be associated with an increased risk of infection.

The estimates of the overall FT contained significant scatter and some clear outliers in keeping with highly variable data (Figure 2). The largest FT was found in the case of the non-mechanically ventilated environment with windows closed throughout, where in some cases the concentration levels had not returned to baseline after more than an hour. The estimates for the required FT were notably smaller for the procedures in the hospital mechanically ventilated closed and open bays. With two exceptions, the aerosol levels were found to return to pre-procedure levels within less than 10 minutes. This demonstrated the effectiveness of the ventilation system at reducing the required FTs, at least when operated at 6 ACH.

FT estimates were larger for the procedures in which the tooth being operated on was alternated every 5 minutes (Figure 2), indicating that even with mechanical ventilation, the FT required was dependent on the procedure being carried out.

## Discussion

There is no published research evaluating aerosol procedures within dentistry which directly relates to contemporary practice in both hospital and private practice setting. This is of particular importance with the current return to practice initiatives across the world and the development of guidelines for dentists. Previously published studies represent the behaviour of droplets (splatter) in the dental clinics after AGP, rather than considering aerosol (of <5μm) (Shahdad et al. 2020). Moreover, a recent literature review concluded that the current evidence base cannot support a defined and appropriate FT for dental AGPs in the context of the COVID-19 pandemic with very weak evidence that peaks in bacterial dissemination during dental procedures which may take approximately 30 minutes to dissipate (SDCEP 2020; Innes et al. 2021).

Our findings indicate that a key arbiter of the delivery of safe dental care during Covid-19 is ventilation. We noted a marked decrease in aerosol in the locality with the use of mechanical HEPA filtered mechanical ventilation, opening of windows and the use of EOS. The impact of ventilation was best represented during the procedures in practice with non-mechanical ventilation when windows were left closed throughout with an increase in the concentration of aerosols across all size ranges during the procedure followed by a slow decline during the FT. In some instances, the FT exceeded 90 minutes and therefore, our results strongly indicate that AGP’s should not be carried out in surgeries without ventilation and corroborates the findings of a recently published scoping review (SDCEP 2020). Figure 2 (a, iii) demonstrates the risk of undertaking AGPs in a non-ventilated surgery. The procedures were carried out back-to-back without any air change between procedures and with each subsequent experiment the level of concentration increased. In practice this would translate to an increasing risk of transmission to patient and staff after every AGP.

When windows were left open after the procedure, effective exchange between the indoor and outdoor air was observed, and the increase in concentration would not represent an increase risk of transmission, as the outside air is most likely a representative of outdoor pollutants (Gouriou et al. 2004). This suggests effective dilution of aerosol by natural ventilation after opening a window at the end of an AGP. Natural ventilation, such as a window allows outside air to mix with room air to dilute any aerosol; however, it is claimed that it is not possible to quantify the number of ACH due to variation in effectiveness of dilution and so would impact on calculation of FT (SDCEP 2020). Theoretical modelling of airborne contaminants has been reported to predict FT at a wide range of air change rates, for AGPs of varying lengths, and with or without procedural mitigation (SDCEP 2020). The modelling (FGDP 2020) makes a number of assumptions, including that all procedures would generate aerosolised virus at the same rate and that aerosols and larger droplets produced by dental procedures will only be removed by dilution. The accuracy and validity of such tools is difficult to ascertain as the algorithm and data used for calculation is unclear and not publicly available.

In this study, mechanical ventilation with six air exchanges per hour, such as those in most modern hospital environments, showed low pre-procedural particulate counts at all particle sizes. Modern systems appear to require lower FT due to their efficiency and more importantly reduced risk to operators and those in the nearby surroundings. This study corroborated the findings of a previous study in which splatter from AGP’s did not show any difference in distribution between open clinic and closed surgery environments (Shahdad et al. 2020). Although there were small spikes recorded in the adjacent bay, the increases tended to be broad (i.e. not dominated by isolated spikes) and occurred 10-20 minutes after the start of the procedure. These were not repeatable and seemed random, so would be considered less relevant. These effects were likely to be a result of gradual dispersion of aerosol from the patient. Diffusion homogenises the concentration of particles leading to a reduction in the magnitude of spikes which was confirmed in that the emissions from the procedures themselves far exceed those from any other non-dental source.

Mitigation has been reported in national and international guidance’s and documents produced by working groups as a method of reducing aerosol. Most guidance’s are, however, largely based on outdated research or data pertaining to splatter rather than true aerosol (Shahdad et al. 2020). The NSS technical report indicated that 10 minutes was necessary to allow droplets (>5-10 μm) to settle, regardless of air change rate, and that standard infection control precautions, which are well rehearsed in dental practice, are sufficient to mitigate the hazard (National Services Scotland Short Life Working Group 2020). Previously, the use of EOS has demonstrated reduction of splatter (intensity of contamination and frequency) during most of the simulated procedures in both open clinic bays and closed bays in the same mechanically ventilated setting as used in this study (Din et al. 2020; Shahdad et al. 2020). In another recent study, EOS was found to statistically significantly reduce the aerosol particulate levels during various AGPs (Nulty et al. 2020).

In this study, the additive effect of EOS on aerosol in reducing the FT in non-mechanically ventilated environment was confirmed. This was unsurprising given the lack of any mechanical ventilation other than the EOS. However, the additive effect of EOS was less noticeable in the hospital environment involving HEPA-filtered mechanical ventilation implying that the ventilation system in isolation was potent. EOS was less likely to reduce the median overall FT in an open bay environment. This reflects the fact that in a closed bay, the effect of the EOS on the overall ACH was larger than in the open bay characterised by a larger area and a greater number of ventilation ports. Notwithstanding this, the particle spikes recorded during the AGP were less noticeable in both open and closed bays when using EOS and without an EOS, these small spikes represented the majority of the increase in aerosol levels.

The data from this study is accurate for the air filtration system within our dental hospital (6 ACH) and variation between filtration systems of different makes, models and age may be more or less effective than the system tested.

Given the rapid evaporation of very small droplets, the aerosols can be expected to comprise primarily of solid droplet nuclei such as tiny fragments of the tooth (Xie et al. 2007). The production of these fragments is likely to depend on the physical properties of the tooth, any restorative material and the cutting action at the chipping interface. Undertaking simulation on extracted natural teeth under the same conditions as the plastic manikin teeth identified a lack of variation in particle characteristics, and therefore, the results of this study can be interpreted with confidence as being representative of a real-world scenario.

Dentists have access to a large variety of dental equipment, and there will undeniably be variation in the aerosol production between these products and their mode of use. For example, not only would we expect differences in the amount of water coolant released during fast handpiece use but also the force at which this is released and the subsequent spread of this into the environment. FT estimates were larger for the procedures in which the tooth being drilled was alternated every 5 minutes. This suggested that the position and strength of suction is vital in reducing aerosol levels, and optimal techniques are recommended whereby suction is placed at the operating site prior to the operation of the FHP, maintaining an intimate relationship when moving between operating sites. Equally, it would be sensible to stop the hand piece before changing position to reduce the escape of aerosol from the suction inlets.

Furthermore, as with all experiments of this nature, all results are specific to the operators and assistants conducting the procedures. As such, there will undoubtedly be variations in aerosol production associated with different operators and assistants. The procedures were conducted using a manikin thereby eliminating patient factors such as movement, saliva, tongue and involuntary actions, which may have an impact on the amount of saliva generated in real patients. The authors suggest that future research specifically look at the aerosolization of respiratory viruses and the associated fallow times.

## Conclusions

Within the limitations of this study, the following conclusions can be drawn:

1. Ventilation in dental practices is an essential pre-requisite for carrying out AGPs; no AGP should be carried in rooms without ventilation.
2. Recommended FT in a HEPA filtered mechanically ventilated room with at least 6 ACH may be as little as 10 minutes.
3. The EOS system reduced the peaks in particle concentration in non-mechanically ventilated and mechanically ventilated environments.
4. Careful four-handed dentistry with HVS and SE appears to remain the primary mitigating method.

## Data Availability

Raw data is available by contacting the authors

## Acknowledgements

TSI Incorporated, USA. Testing equipment was provided by TSI Incorporated and their involvement with this study was on an advisory basis only and had no bearing to the outcomes achieved. Specialist Dental Services, Harley Street, UK. For allowing us access to a primary care setting for non-mechanically ventilated surgeries representative of most dental provision in the UK.

